# Clinical and behavioural characteristics of self-isolating healthcare workers during the COVID-19 pandemic: a single-centre observational study

**DOI:** 10.1101/2020.05.07.20094177

**Authors:** Angus de Wilton, Eliz Kilich, Zain Chaudhry, Lucy CK Bell, Joshua Gahir, Jane Cadman, Robert A Lever, Sarah A Logan

## Abstract

**Objectives:** To describe a cohort of self-isolating healthcare workers (HCWs) with presumed COVID-19.

**Design:** A cross-sectional, single-centre study.

**Setting:** A large, teaching hospital based in Central London with tertiary infection services.

**Participants:** 236 HCWs completed a survey distributed by internal staff email bulletin. 167 were female and 65

**Measures:** Information on symptomatology, exposures and health-seeking behaviour were collected from participants by self-report.

**Results:** The 236 respondents reported illness compatible with COVID-19 and there was an increase in illness reporting during March 2020. Diagnostic swabs were not routinely performed.. Cough (n=179, 75.8%), fever (n=138, 58.5%), breathlessness (n=84, 35.6%) were reported. Anosmia was reported in 42.2%. Fever generally settled within 1 week (n=110, 88%). Several respondents remained at home and did not seek formal medical attention despite reporting severe breathlessness and measuring hypoxia (n=5/9, 55.6%). 2 patients required hospital admission but recovered following oxygen therapy. 84 respondents (41.2%) required greater than the obligated 7 days off work and 9 required greater than 3 weeks off.

**Conclusion:** There was a significant increase in staff reporting illness compatible with possible COVID-19 during March 2020. Conclusions cannot be drawn about exact numbers of confirmed cases due to lack of diagnostic swabbing. There were significant numbers of respondents reporting anosmia; as well as early non-specific illness prior to onset of cough and fever. This may represent pre-symptomatic HCWs who are likely to be infectious and thus criteria for isolation and swabbing should be broadened. The study also revealed concerning lack of healthcare seeking in respondents with significant red flag symptoms (severe breathlessness, hypoxia). This should be addressed urgently to reduce risk of severe disease being detected late. Finally, this study should inform trusts that HCWs may require longer than 7 days off work to recover from illness.

Strengths and limitations of this study
- To the authors’ knowledge, this study presents one of the first descriptive data analysis of self-reported healthcare worker (HCW) COVID-19 exposures and symptomatology in the UK.
- Study respondents represented a broad range of job roles, including both frontline clinical and non-patient facing staff.
- The inclusion of questions focusing on health-seeking behaviour allows results to be used to inform human resource management in the developing pandemic, and provides concerning but important data around late healthcare seeking in HCWs
- Data were self-reported, cross-sectional and retrospective, which may be subject to recall bias, and the lack of diagnostic swabbing in the majority of respondents limits interpretation of the data
- Full demographic data were not collected on participants and certain staff groups may have been over-represented in the sample, which may introduce sampling bias.

## BACKGROUND

In response to the COVID-19 pandemic, the UK government enacted a range of policies in an attempt to limit the spread of infection. These included guidelines on self-isolation for symptomatic individuals and a formal social distancing policy [1]. Healthcare workers (HCW) are at a disproportionate risk for COVID-19 disease through occupational exposure. Additionally, there are emerging concerns that HCWs may be at an additional risk of developing severe disease through repeated exposure to high viral load in the clinical environment [2], This has implications for workforce planning and operational preparedness in the current crisis.

Testing for SARS-CoV-2 has not been routinely available for UK healthcare workers. Instead, HCWs have been advised to self-isolate for a minimum of 7 days from the onset of symptoms and return to work after this period if symptom free [3]. Earlier guidance (during the period of survey completion) advised seeking formal medical attention if difficult breathing developed, this was later updated to any subjective deterioration or failure to improve [4], Avenues of medical advice open to patients in the UK include: NHS 111, a free online and telephone triage and advice system; the patient’s own general practitioner (GP); and Accident and Emergency (A&E) departments in secondary care. These resources are in addition to informal health advice provided by friends, family or colleagues. Little is currently described about healthcare seeking behaviour in HCWs.

A range of symptoms have been described in individuals with suspected and confirmed COVID-19, including fever, cough and shortness of breath [5,6]. Despite a multitude of recent clinical studies there is still limited published work describing symptoms amongst HCWs [7,8]. Given the differential occupational exposure of HCWs, there is the potential that the spectrum of symptoms experienced may differ from that of the general public.

In this study we describe the experiences and symptoms of HCWs self-isolating for presumed COVID-19 infection in a cohort of frontline HCWs in a central London teaching hospital. We also characterise behaviours surrounding self-care, isolation and return to work. This will aid in better understanding the spectrum of illness amongst HCWs, inform future HCW testing strategies and provide data to support human resource management in the developing pandemic.

## METHODS

We performed a cross sectional, single-centre study of NHS HCWs who had self-isolated with symptoms compatible with COVID-19 prior to April 2020. Participants were recruited from University College London Hospitals NHS Foundation Trust by distributing the voluntary survey via staff email bulletin to all staff departments from 1st April 2020 - 10th April 2020. Consent was obtained via electronic signature. Responses were devoid of personal identifiers and were collected and processed via Form Assembly Enterprise cloud https://www.formassemblv.com/. All data was stored in compliance with the General Data Protection Regulation (EU GDPR 2016/679) and Data Protection Act (UK 2018). The Study was approved by the Audit and Research Committee at the Hospital of Tropical Diseases, UCLH [9], Anonymous data was exported to Microsoft Excel 2010 (Microsoft Corporation) and R (R Development Core Team 2008) for analysis. Venn diagrams were generated using Venny 2.1.0 (https://bioinfoep.cnb.csic.es/tools/vennv/) [10] and BioVenn (https://www.biovenn.nl/) [11],

## RESULTS

### Demographics, timeline and exposure history

There were 236 respondents to the survey of which 167 (70.8%) were female and 65 (27.5%) male (Table 1). Respondents were aged between 18 and 71. The respondents were from a broad range of hospital roles with the most common groups being doctors (33.5%) and nurses (25.4%). There was a broad range of other professions represented. The majority were non smokers (79.7%), with 32 (13.6%) ex-smokers and eight current smokers (3.4%). Twenty-four respondents (10.2%) reported being in a vulnerable group according to Public Health England criteria [1].

**Table 1:** Demographics of respondents. Demographic data collected via survey from staff at University College Hospital London.

| Demographic | n=236 (%) |
| --- | --- |
| Sex |  |
| Female | 167 (70.8) |
| Age |  |
| 18-28 | 53 (22.5) |
| 29-39 | 88 (37.3) |
| 40-50 | 66 (28.0) |
| 50-60 | 21 (8.9) |
| 61-71 | 3 (1.3) |
| **Workplace** |  |
| UCH* | 229 (97.0) |
| Other | 4 (1.7) |
| Hospital | 228 (96.6) |
| Community | 5 (2.1) |
| **Job Role** |  |
| Doctor | 79 (33.5) |
| Nurse | 60 (25.4) |
| Administrator | 18 (7.6) |
| Other | 17 (7.2) |
| Other allied healthcare professional | 17 (7.2) |
| Radiographer | 14 (5.9) |
| Manager | 9 (3.8) |
| Healthcare assistant | 8 (3.4) |
| Physiotherapist | 6 (2.5) |
| Dietician | 4 (1.7) |
| Other non-clinical support | 2 (0.8) |
| Occupational Therapist | 1 (0.4) |
| **Smoking Status** |  |
| Smoker | 8 (3.4) |
| Non-smoker | 188 (79.7) |
| Ex-smoker | 32 (13.6) |
| **Vulnerable Group†** |  |
| Yes | 24 (10.2) |

\*University College Hospital
†As defined by Public Health England
[https://www.gov.uk/government/publications/covid-19-guidance-on-social-distancing-and-for-vulnerable-people/]

Known direct contact with SARS-CoV-2 positive patients was reported in 81 HCWs (34.3%) (Figure 1A), of which 24 (29.1%) of these were in appropriate personal protective equipment as perceived by respondents. Over half of those surveyed (128 respondents, 53.4%) were not aware of having had any direct contact with COVID-19 positive patients. Initial suspected cases were identified in mid-February and increased until late March 2020 (Figure 1B).

**Figure 1:**
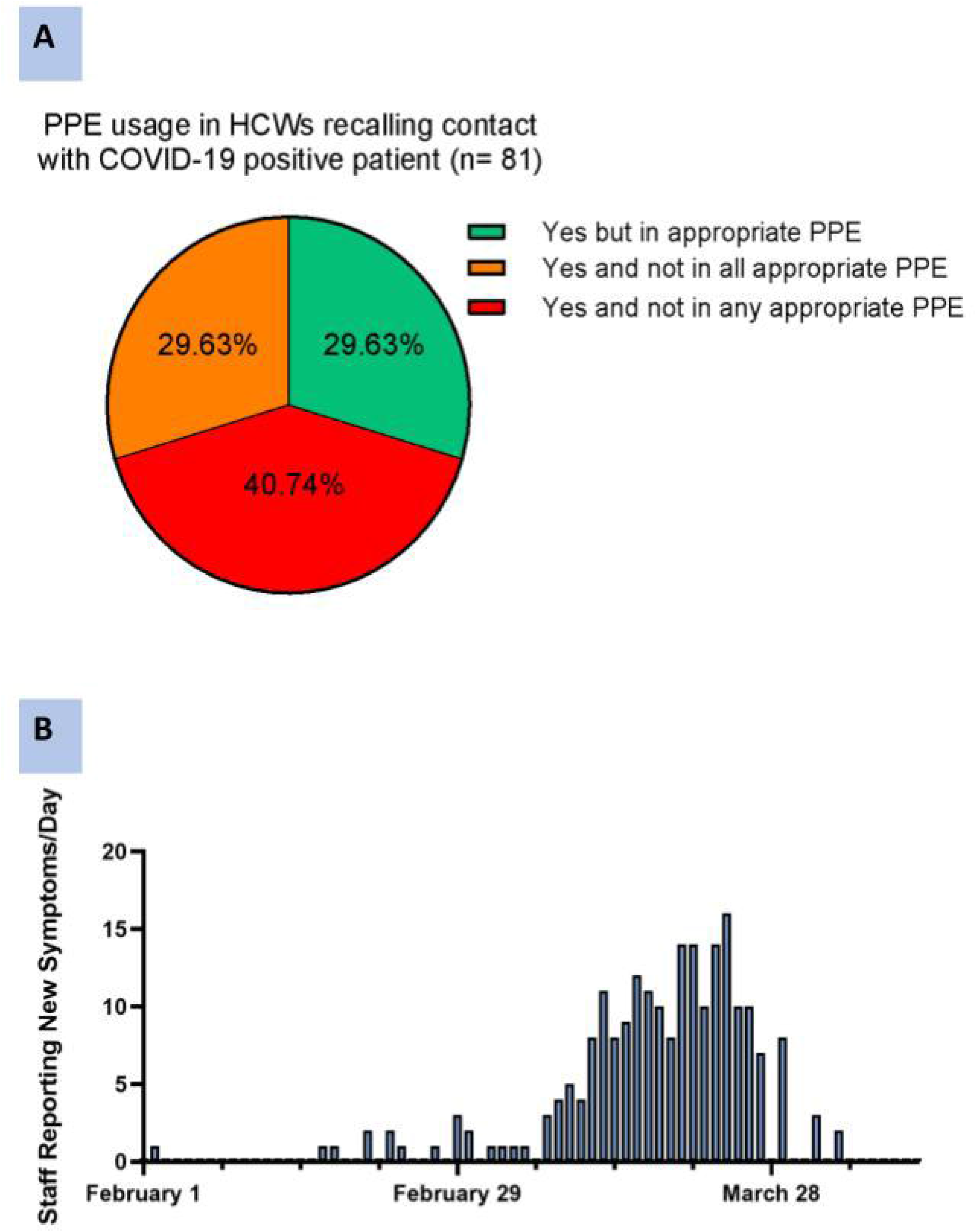
Self-perceived PPE usage and date of symptom onset In healthcare workers. **(a)** 81 of 236 respondents reported an exposure to a patient who was confirmed or subsequently confirmed to be SARS-CoV-2 positive. The pie chart shows the breakdown of responses in this group when asked whether they considered that they were wearing appropriate PPE, partly appropriate PPE or no appropriate PPE at the time of this exposure. 40.74% of respondents in this group (n=33, 13.98% of overall cohort) reported they considered that they were not wearing any appropriate PPE at the time of exposure, **(b)** Respondents were asked to report their first day of symptom onset. Most reported symptom onset occurring within the first 3 weeks of March 2020.

### Clinical symptoms reported during self isolation

The most commonly reported symptoms included headache (78.8%), cough (75.8%), myalgia (63.6%) and fever (58.5%) (Table 2). Eighty-four respondents (35.6%) reported dyspnea during their illness; of these 41 patients (17.4%) reported shortness of breath only on exertion with only 12 patients (5.1%) reporting shortness of breath at rest. Nearly one third of HCWs experienced symptoms beyond 14 days (73 respondents, 30.9%).

**Table 2:**
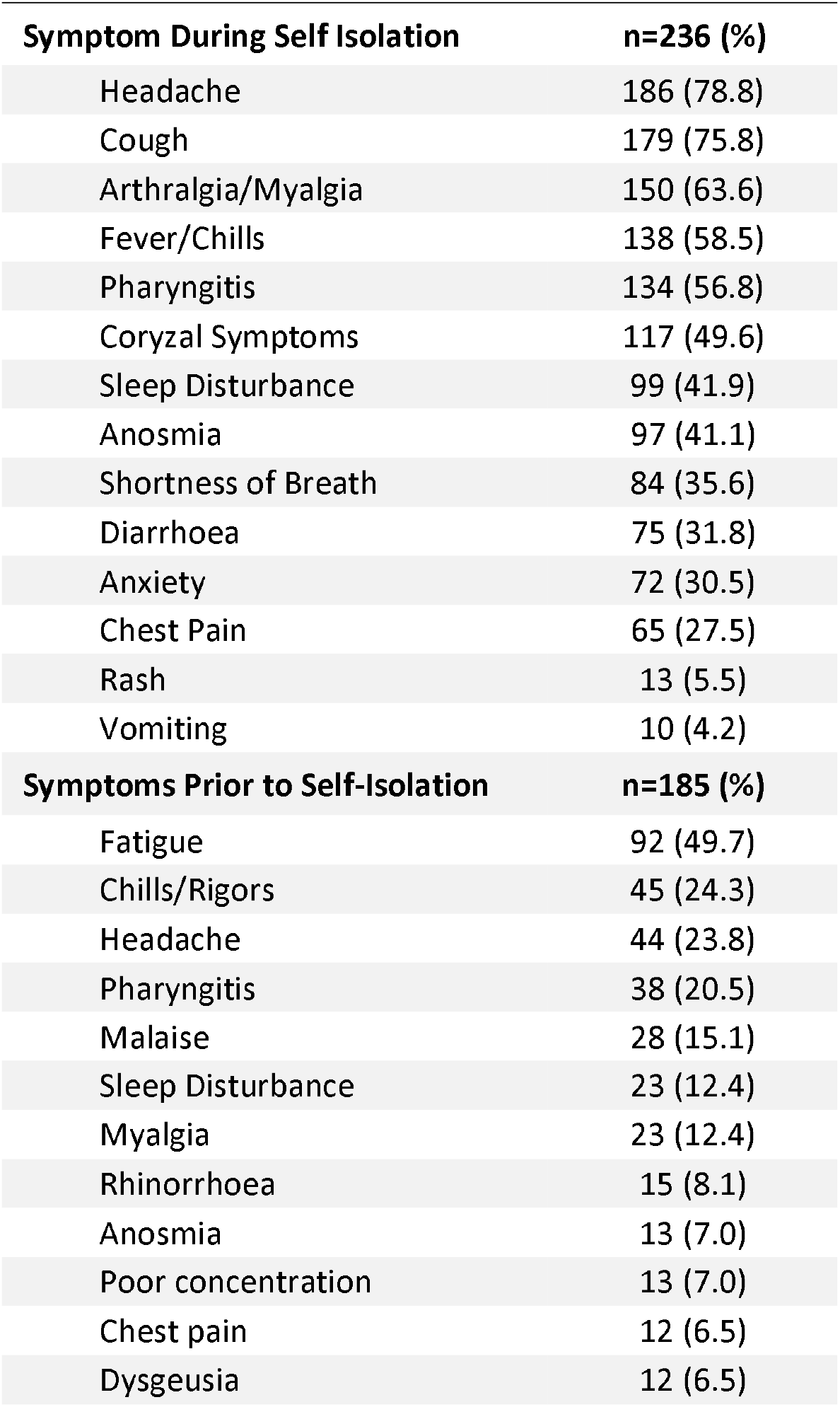
Symptoms during self isolation; symptoms prior to self isolation. Respondents were asked to report symptoms they experienced whilst self isolating and unwell with an illness they perceived to be COVID-19. The number of respondents reporting each symptom is shown. Respondents were subsequently asked to report symptoms that preceded self isolation. The symptoms occurring earlier in their illness prior to self isolation are shown in the lower section of the table.

| Symptom During Self Isolation | n=236 (%) |
| --- | --- |
| Headache | 186 (78.8) |
| Cough | 179 (75.8) |
| Arthralgia/Myalgia | 150 (63.6) |
| Fever/Chills | 138 (58.5) |
| Pharyngitis | 134 (56.8) |
| Coryzal Symptoms | 117 (49.6) |
| Sleep Disturbance | 99 (41.9) |
| Anosmia | 97 (41.1) |
| Shortness of Breath | 84 (35.6) |
| Diarrhoea | 75 (31.8) |
| Anxiety | 72 (30.5) |
| Chest Pain | 65 (27.5) |
| Rash | 13 (5.5) |
| Vomiting | 10 (4.2) |
| Symptoms Prior to Self-Isolation | n=185 (%) |
| Fatigue | 92 (49.7) |
| Chills/Rigors | 45 (24.3) |
| Headache | 44 (23.8) |
| Pharyngitis | 38 (20.5) |
| Malaise | 28 (15.1) |
| Sleep Disturbance | 23 (12.4) |
| Myalgia | 23 (12.4) |
| Rhinorrhoea | 15 (8.1) |
| Anosmia | 13 (7.0) |
| Poor concentration | 13 (7.0) |
| Chest pain | 12 (6.5) |
| Dysgeusia | 12 (6.5) |

A number of those surveyed reported a prodromal syndrome prior to the appearance of the symptom which precipitated self-isolation. These included fatigue (92 respondents, 39.0%), chills (45 respondents, 24.3%) and headache (44 respondents, 23.8%).

### Dynamics and clustering of symptoms reported during illness

Of 125 HCW with fever only 15 (12%) reported fevers beyond day 7 of illness (Figure 2A). Most respondents reported at least one of the most commonly reported three symptoms with only 9 individuals (3.8%) not reporting headache, cough and/or myalgia (Figure 2B). Assessing symptom overlap demonstrated that for the three most commonly reported symptoms in the cohort (headache, cough & myalgia), approximately two-thirds of respondents reporting each symptom also reported fever (Figure 2C). Correlation analysis of all symptoms did not demonstrate any clear symptom clusters except from cough, shortness of breath and chest pain. Shortness of breath and chest pain were described by a minority of respondents (Figure 2D).

**Figure 2:**
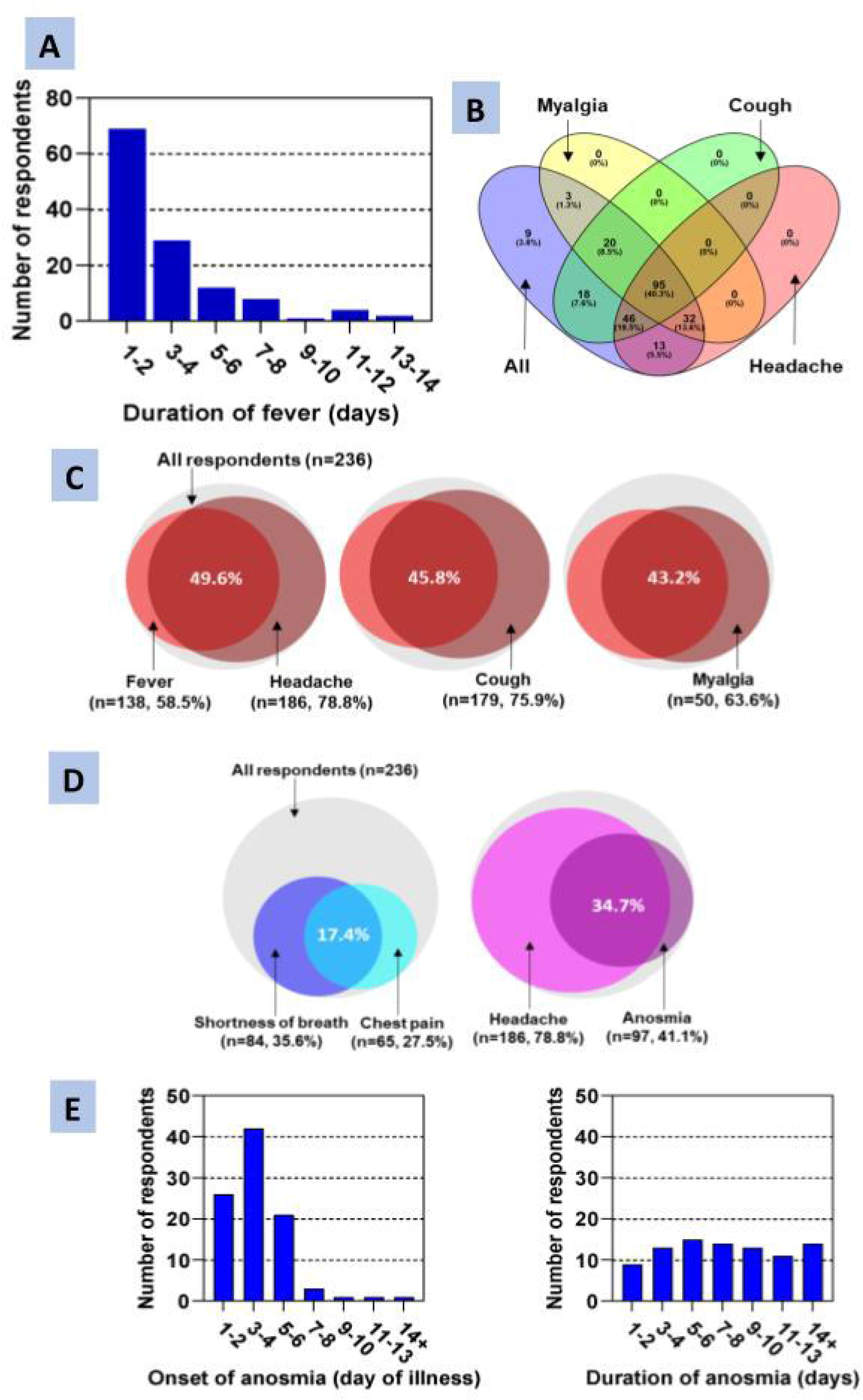
Duration of reported fever In self isolating healthcare workers (A); symptom clustering reported during illness (B, C, D); and characterisation of anosmia (E) **(a)** respondents were asked to report the duration of their fever. The majority of respondents reported fever duration less than 7 days (n=110, 88). Fever persisted to 7 days or more in 12% (n=15) **(b)** Non-proportional Venn diagram (generated using Venny^1^) demonstrating the crossover between the three most commonly reported symptoms (headache, cough and myalgia). The purple ellipse demonstrates the all patient denominator. Only 9 respondents (3.8%) did not report any of these symptoms, **(c)** Proportional Venn diagrams (generated using BioVenn^2^) demonstrating the crossover between fever (reported by 58.5% of respondents) and the three most commonly reported symptoms – headache, cough and myalgia. Grey circles demonstrate the denominator (all respondents), light red circles show respondents reporting fever and burgundy circles show respondent reporting other symptoms. Percentages in white show the proportion of the overall group of respondents reporting both fever and the relevant second symptom.**(d)** Proportional Venn diagrams (generated using BioVenn^2^) demonstrating the crossover between shortness of breath & chest pain, and headache & anosmia. Grey circles demonstrate the denominator (all respondents). Percentages in white show the proportion of the overall group of respondents reporting both symptoms in each Venn, **(e)** Respondents reporting anosmia (n=91, 41.1%) were asked the day of onset and duration of this symptom. The majority of respondents developed anosmia early in illness (median day 3, SD 1.96) and had resolution of anosmia within 2 weeks of its onset (n=75, 84%).

A variety of neurological syndromes have been linked to COVID-19 [11], In particular anosmia has been reported to be a specific indicator of COVID-19 disease [12]. Ninety-one respondents described anosmia during their illness (41.1%) and 13 reported anosmia as part of a viral prodrome (7.0%). Most individuals with anosmia also reported headache, although this was in the context of headache being the most common symptom overall (78.8% of respondents) (Figure 2D). Onset of anosmia peaked at day 3-4 of illness with 84% reporting symptomatic resolution within 14 days (Figure 2E).

### Healthcare seeking behaviour in HCWs

Respondents commonly sought advice regarding their illness informally (26.7%), via NHS 111 (25.4%) and general practice (7.6%). Two patients attended Accident and Emergency and required oxygen therapy during hospital admission. No respondents required non-invasive or invasive ventilation.

A minority of respondents had access to monitoring of oxygen saturations during their illness, and of those who commented on saturation levels, 11 (4.7%) described saturations below 94% during their illness to variable degrees of desaturation. Two respondents reported saturations below 85% either at rest or exertion. Only 41.7% of those with breathlessness at rest (n=12) and 44.2% of those who were breathless on exertion (n=43), sought formal medical advice (Figure 3A). Notably 9 respondents reported a combination of breathlessness and saturations less than 94% at rest; of these respondents 5 (55.6%) did not seek any formal healthcare advice (Figure 3B).

**Figure 3:**
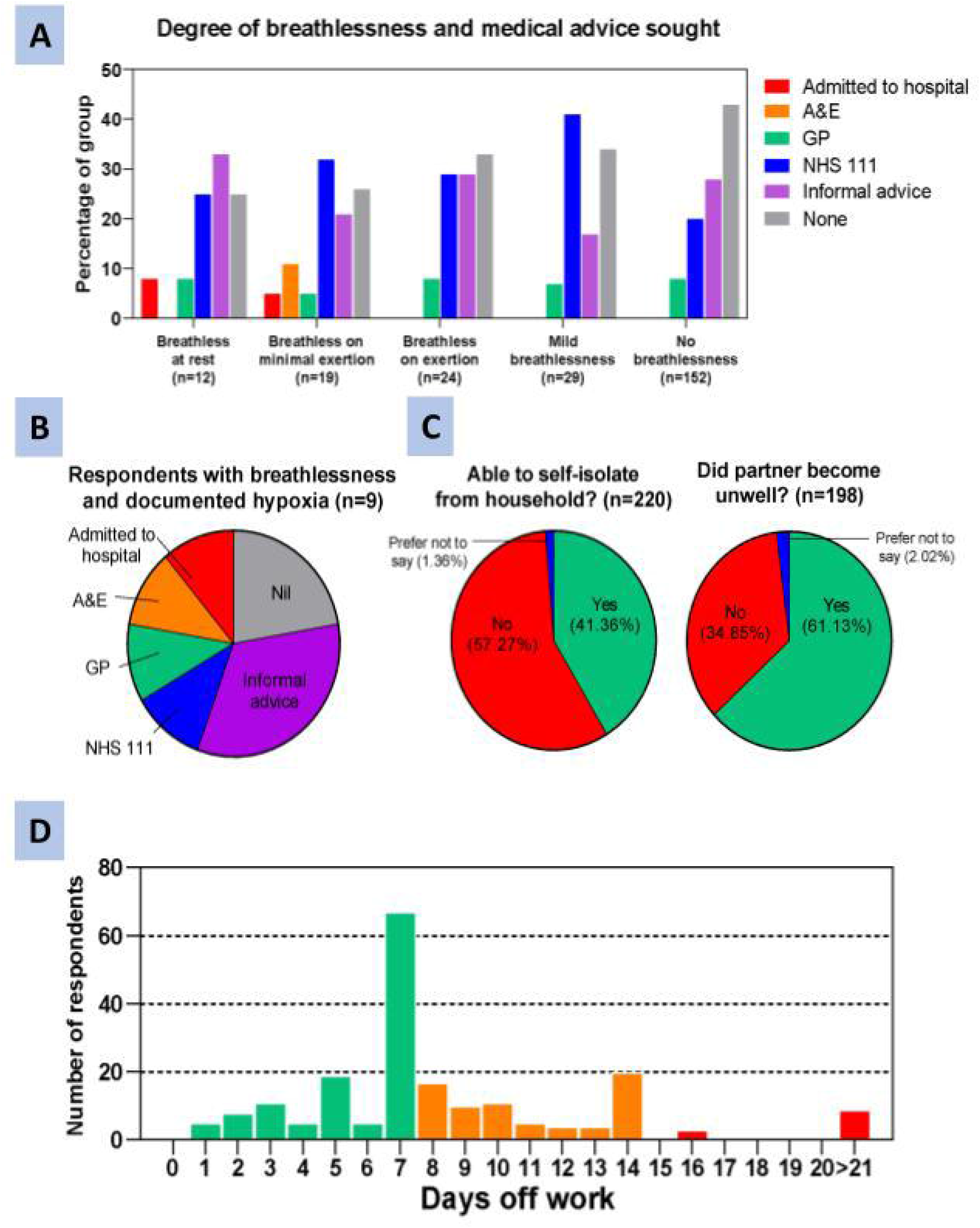
Healthcare seeking behaviour as triggered by breathlessness in HCWs (A); Access to self isolation facilities (B); illness in close contacts of HCWs (C); return to work timeline (D) (a) 84 respondents reported breathlessness (35.6%); increased severity of breathless did not appear to lead to increased formal healthcare seeking in respondents. Of those respondents reporting breathlessness at rest (n=12), only 41.7% (n=5/12), sought formal medical attention (NHS 111, GP, A&E) (b) 9 respondents reported a combination of breathlessness and saturations of <94% (measured using home oximeters). A majority (n=5/9,) of those respondents sought either no or informal advice only, (c) Respondents were asked if they felt able to self isolate away from other household members (seperate bedroom, bathroom). A majority did not feel able to to self isolate in this way (n= 126, 57.27%). (d) Respondents were asked whether their partner became unwell (phrased as ‘sharing bed on night of symptom onset’) during 14 days after symptom onset. A majority (n=125, 61.13%) reported their partners did become unwell during this period.

### Self isolation and return to work

The majority of respondents (57.3%) did not feel able to effectively distance themselves from household members whilst unwell (as defined as access to a separate bedroom and/or toilet) (Figure 3C). Close contacts (defined as sharing a bed with symptomatic respondent on night prior to symptom onset) frequently became unwell during the 14 days after symptom onset of respondents (Figure 3C). Time to return to work varied in this cohort, with a significant number of respondents requiring greater than 7 days off work prior to return (Figure 3D). Nine respondents required greater than 3 weeks off work (4.4%). In addition 20% of respondents felt they returned to work before they felt ready.

## DISCUSSION

To the authors knowledge this is one of the first descriptive studies on the presentation of Presumed COVID-19 in HCWs in the UK. The only other peer reviewed study looking at HCW COVID-19 infection in the UK by Hunter et al [13] reported SARS-CoV2 positivity rates during a HCWs testing programme in March 2020. They found a steep rise in confirmed COVID-19 cases corresponding with the data we present on presumed infections. Whilst an important insight into the epidemiology of COVID-19 in HCWs, this study did not provide data on clinical presentation or behavioural aspects of HCW infection, and occupational data was not available in the majority of participants. This study therefore provides novel data on healthcare seeking behaviour, self-isolation facilities, and return to work timelines of HCWs with presumed COVID-19. The study is limited by the fact that respondents did not have access to diagnostic swabs during their illness and therefore any non-COVID-19 related symptoms triggering self-isolation (e.g. due to other respiratory viral infections) amongst respondents may confound conclusions derived from these results. Whilst the data should therefore be interpreted with caution, the overall data temporally match with the COVID-19 outbreak across London. This suggests that self-reported symptoms may be a reasonable surrogate for the illness during the outbreak. We identified 2.9% of the 9000 clinical and administrative staff members at UCLH reporting symptoms consistent with COVID-19 [14]. The use of a voluntary online survey to collect this data has a number of sources of bias. In particular it is unlikely that every self-isolating HCW was captured by this approach and we suspect that our sample over-represents the number of training grade doctors in our sample given that 33.9% of the survey population were doctors. This could represent that clinical facing staff (nurses, doctors and healthcare assistants) were more likely to develop symptoms. Alternatively, they could have been more likely to respond to the survey given the email bulletin tends to reflect clinical guideline changes and up to date policy each day. The peak of cases seen at the end of March 2020 may indicate increased community transmission, with lockdown being introduced in the UK on 23rd March 2020 and after social distancing measures. It could also represent staff-to-staff transmission prior to the guidance from Public Health England to self-isolate with symptoms. Future studies could evaluate contact tracing in relation to staff-staff interactions during a lockdown period when R0 is believed to be <1 for the community population to estimate nosocomial spread.

The symptoms described by respondents are in keeping with those described in previous cohorts of non severe COVID-19 patients. In our study there was a significant proportion of this cohort reporting anosmia - 41.5% - which has not been described to this extent in other upper respiratory tract infections [15]. We also describe respondents reporting non-specific symptoms early in their illnesses prior to onset of self isolation triggered by cough or fever. The presence of sore throat, profound fatigue and anosmia early in the presentation should therefore prompt staff testing strategies to include all these symptoms in the clinical case definition even in the absence of fever or new continuous cough. A diagnostic swab early on in the course of the syndrome will enable health care workers who are infectious to stay at home to limit onward transmission within healthcare settings.

Our findings indicate that for the majority of healthcare workers this is a self-limiting illness of less than one week. However, a significant minority remain unwell beyond 8 days and many have protracted illnesses beyond this. Notably, severe COVID-19 illness in HCW is likely to be under-represented in our sample due to selection bias. A particular point of concern raised by our study is that a significant proportion of healthcare workers did not seek formal medical advice or assessment despite significant shortness of breath and/or hypoxia. More worrying perhaps also is the HCWs who reported significant breathlessness and measured hypoxia using home oximetry and still did not seek formal medical attention. The reasons for this reluctance to seek formal care were not captured by the survey. It will be important for Trusts across the United Kingdom to ensure they have mechanisms in place to encourage staff to seek medical advice if they develop shortness of breath, and ensure adequate pathways are in place to support such seeking of care. This may be an important point of intervention to reduce HCW morbidity and mortality.

Many staff reported symptoms prior to 13th March 2020, after which Public Health England introduced guidance to healthcare workers to self-isolate for 7 days if they had symptoms of a continuous cough or fever (Figure 1). These individuals were not subject to isolation restrictions and therefore many of whom isolated for under 7 days or not at all. Our findings indicate that HCW respondents lack the ability to self isolate effectively away from family members, and that significant numbers of close contacts of HCWs became unwell in the period following respondents’ symptoms. Hospital trusts should consider provision of hospital accommodation for staff unwell with COVID-19 to minimise onward transmission to their home contacts; especially when household contacts of HCWs are in vulnerable groups. The return to work data in this study suggests that hospital trusts should expect significant numbers of HCWs to require beyond 7 days of leave prior to return. Current guidelines to self isolate for 7 days may lead to an expectation of return to work at this stage, whereas our data suggests many staff will require more time off work than the minimum recommended by PHE. Indeed, 1 in 5 staff felt they returned to work too soon. Staff members will therefore need support to ensure they are back to full health prior to returning to work and feel supported in this return and this could help inform occupational health and workforce management policy going forward.

In summary this study was limited because the majority of respondents were not confirmed with positive swabs. Given the lack of access to a diagnostic test for the majority of respondents the survey may include participants with illnesses that mirror COVID-19. In addition, we may have missed recruiting participants who had very mild or very severe symptoms that may not have completed the survey. Recruitment occurred by email bulletin which may lend bias to response rates in different staff groups, particularly clinical and patient facing staff who may be more likely to check bulletins for daily guidance changes. Future studies looking at symptom prevalence, healthcare seeking behaviour and return to work in HCWs in confirmed cases of COVID-19 will further inform the area. Similarity in data between our study and confirmed cohorts may validate the use of symptom surveys in data collection for staff illness in COVID-19. Hospital trusts should urgently address issues raised regarding delayed healthcare seeking in staff with hypoxia and severe breathlessness.

## Data Availability

Data available on reasonable request to responses will be within 28 days

## Acknowledgements

We thank Form Assembly https://www.formassemblv.com/ for the generosity In providing the Enterprise Cloud Account to facilitate research during the COVID-19 pandemic.

## Contributorship statement

**<u>ADW and SL conceived of the presented idea. All authors contributed to study design, conduct and data analysis. ADW led writing the manuscript. All authors contributed to and provided critical feedback to the manuscript. The corresponding author attests that all listed authors meet authorship criteria and that no others meeting the criteria have been omitted.</u>**

